# PREVALENCE, PATTERN, AND CORRELATES OF DEPRESSION AMONG DRUG-SUSCEPTIBLE TUBERCULOSIS PATIENT ENROLLEES IN OGBOMOSO, OYO STATE: A CROSS-SECTIONAL STUDY

**DOI:** 10.1101/2025.02.26.25322944

**Authors:** Sunday Olakunle Olarewaju, Sunday Charles Adeyemo, Ayomide Timilehin Kayode, Janet Oluwaseyi Ayinmodu, Olushina Ololade Oladeji, John Temitayo Odedele, Doyin Victoria Olaniyan, Oluwatoba Jeremiah Oyedeji, Zainab Adedamola Abdulsalam, Eniola Dorcas Olabode, Ayodele Raphael Ajayi

## Abstract

Tuberculosis (TB), an infectious disease caused by *Mycobacterium tuberculosis* is still one of the leading public health problems, despite advances in the effort to reduce its incidence, morbidity, and mortality. Studies have shown that the prevalence of depression correlated with the severity and duration of tuberculosis. Therefore this study aims to find out the prevalence and pattern of depression among drug-susceptible TB patients to improve treatment outcomes and thereby reduce morbidity and mortality from the disease.

The study was a cross-sectional hospital-based survey. Sample size of 333 respondents was calculated using Leslie Fischer’s formula (n= z^2^pq /d^2^). A multistage sampling technique was used to select respondents. Data was collected using a pre-tested semi-structured questionnaire and analyzed using Statistical Package for Social Sciences (SPSS) version 20. Descriptive analysis was done or all variables. Bivariate and multivariate analysis was done using chi-square and binomial regression respectively. The level of significance is set with a p-value less than 0.05.

More than half of the respondents (186, 55.9%) of the respondents were depressed. The majority (122, 65.5%) of respondents had mild depression, 46 (24.7%) had moderate depression while 18(9.7%) had moderately severe depression. Sex, marital status, level of education and average monthly income were significantly associated with depression status at bivariate level. Multivariate analysis revealed that respondents with no formal education were 6 times less likely to develop depression (AOR = 0.175, P = 0.001). Respondents with Primary level of education were 2 times less likely to develop depression (AOR = 0.427, P = 0.023). Respondents with Secondary level of education were 3 times less likely to develop depression compared to Tertiary level of education. Respondents living with HIV were 35 times more likely to develop depression (AOR = 35.303, P = 0.017) compared to those who were HIV negative.

## INTRODUCTION

Tuberculosis (TB), an infectious disease caused by *Mycobacterium tuberculosis* is still one of the leading public health problems, despite advances in the effort to reduce its incidence, morbidity, and mortality.^1–5^ In 2021, about 10.6 million people were reported by the World Health Organisation (WHO) to be infected with Tuberculosis,^6^ with an estimated 23% of the global burden and 33% of the global TB death in Africa.^7^ Nigeria ranks first in Africa, one of the 10 countries with the highest number of missing TB cases, the sixth in the world, contributing to almost 4.6% of the global burden. It is also responsible for a high triple burden of Drug-susceptible TB; a bacteriologically confirmed or clinically diagnosed case of TB without evidence of infection with strains resistant to rifampicin and isoniazid.^8^

Depression is a mental state characterized by loss of interest, feelings of guilt, disturbed sleep or appetite, loss of self-worth, and usually suicidal thoughts.^1,2^ Chronic pain, frequent hospital admissions, and dependency on the hospital found in patients with Tuberculosis have been reported to be associated with depression.^5,9^ Depression is a common mental disorder, about 5% of adults suffer from depression globally (WHO 2023). Depression is a major cause of suicide with more than 700,000 people dying due to suicide every year.^10^ Studies have shown that the prevalence of depression correlated with the severity and duration of tuberculosis.^11^ Depression that accompanies the disease is often due to the nature of the infection, side effects of medications, and other social determinants of health. Several studies have shown a higher prevalence of depression among patients with Tuberculosis as compared to the general population. A study done in Nigeria by Ige and Lasebikan showed a prevalence of about 45. 5%.^9^

When TB and depression co-exist, patients tend to suffer in silence and when accompanied by poor compliance to medication, the mortality rate also increases. Some previous studies identified a poor degree of suspicion of depression in patients being managed for TB by clinicians,^3,11,12^ therefore, paying attention to some of the psychosocial issues that patients under treatment for tuberculosis experience and the improvement of consultation-liaison psychiatric services may optimise adherence and increase the success of treatment. It is therefore important to find out the prevalence and pattern of depression among drug-susceptible TB patients to improve treatment outcomes and thereby reduce morbidity and mortality from the disease, hence this study.

## METHODOLOGY

This study was carried out in Ogbomoso, Southwestern Nigeria. The study was a cross-sectional hospital-based survey involving quantitative methods of data collection. The study was conducted across the Direct Observed Therapy (DOT) centres in selected local governments in Ogbomoso. The DOT centres owned by both state and local governments were used for the study. TB patients, aged 18 years and above, who have been on drugs for two months and there’s proof of drug susceptibility, and who are mentally capable of providing consent were included in the study while newly diagnosed and unregistered pulmonary tuberculosis patients, pregnant women with pulmonary TB, severely ill or debilitated patients, patients with extra-pulmonary TB and patients who cannot give consent were not included. A written consent form was signed by each respondent.

Sample size was calculated using Leslie Fischer’s formula (n= z^2^pq /d^2^) using the proportion for the prevalence of depression among DS TB patients in the previous study which was 27% (0.27).^13^ After 10% non-response was calculated, a total of 333 questionnaires were administered. A multistage sampling technique was used to select respondents; First stage: From the list of urban and rural local government areas in Ogbomoso, Ogbomoso North and Ogooluwa Local Government Areas were chosen.

Second stage: The list of all registered DOT centres in Ogbomoso was collected from the Tuberculosis and Leprosy Supervisor for each Local Government Areas. All DOT centres were included.

Third Stage: Using proportional allocation based on sample size, clients were selected from each DOT centre using a systematic sampling approach until we recruited enough sample allocated percentage.

Data was collected from 25th November, 2024 to 6th December, 2024 using a pre-tested semi-structured questionnaire. Questionnaires were sorted out to check for errors and omissions at the end of the collection of data. Thereafter, data were entered into the computer and analyzed using Statistical Package for Social Sciences (SPSS) version 20. Descriptive analysis was done or all variables. Bivariate and multivariate analysis was done using chi-square and binomial regression respectively. The level of significance was set with a p-value less than 0.05.

## RESULTS

### Sociodemographic characteristics of respondents

Table 1 shows the socio-demographic characteristics of the respondents revealed that the majority, 109 (32.7%), were within the age range of 30–40 years. Most respondents, 216 (64.9%), were married. Regarding education, the largest proportion, 108 (32.4%), had tertiary education. The predominant ethnic group was Yoruba, 271 (81.4%). Most respondents, 174 (52.3%), were Christians.

**Table 1:** Sociodemographic characteristics of respondents.

| **Variables** | **Categories** | **Frequency** | **Percent** |
| --- | --- | --- | --- |
| Age Group | <30 years | 37 | 11.1 |
|  | 30-40 years | 109 | 32.7 |
|  | 41-50 years | 80 | 24.0 |
|  | 51-60 years | 65 | 19.5 |
|  | >60 years | 42 | 12.6 |
|  | Mean±SD = 44.86±14.29 |  |  |
| Sex | Male | 165 | 49.5 |
|  | Female | 168 | 50.5 |
| Religion | Christian | 174 | 52.3 |
|  | Muslim | 147 | 44.1 |
|  | Traditional | 11 | 3.3 |
|  | Others | 1 | 0.3 |
| Marital status | Single | 77 | 23.1 |
|  | Married | 216 | 64.9 |
|  | Widow | 27 | 8.1 |
|  | Widower | 13 | 3.9 |
| Ethnicity | Yoruba | 271 | 81.4 |
|  | Igbo | 21 | 6.3 |
|  | Hausa | 33 | 9.9 |
|  | Others | 8 | 2.4 |
| Level of education | No formal education | 59 | 17.7 |
|  | Primary | 87 | 26.1 |
|  | Secondary | 79 | 23.7 |
|  | Tertiary | 108 | 32.4 |
| Occupational status | Employed | 215 | 64.6 |
|  | Unemployed | 100 | 30.0 |
|  | Student | 18 | 5.4 |
| Average income per | Less than ₦30 000 | 130 | 39.0 |
| month | Above ₦30 000 | 203 | 61.0 |

### Medical history of respondents

Table 2 shows the medical history of the respondents revealed that most respondents, 253 (76.0%), did not have diabetes, and 233 (70.0%) did not have hypertension. Concerning HIV status, the majority, 253 (76.0%), were HIV-negative. A large proportion, 282 (84.7%), had no history of mental illness, and 297 (89.2%) reported no family history of mental illness.

**Table 2:** Medical history of respondents.

| Variables | Categories | Frequency | Percent |
| --- | --- | --- | --- |
| Comorbidities | Diabetes | 49 | 14.7 |
|  | Hypertension | 73 | 21.9 |
|  | HIV | 80 | 24.0 |
| History of mental illness(es) | Yes | 51 | 15.3 |
|  | No | 282 | 84.7 |
| Family history of mental illness(es) | Yes | 36 | 10.8 |
|  | No | 297 | 89.2 |

### Assessment of depression among respondents

More than half of the respondents (186, 55.9%) of the respondents were depressed. Table 3 shows responses of respondents using the PHQ-9. The majority, 168 (50.5%), reported “not at all” for having little interest or pleasure in doing things. Similarly, 168 (50.5%) reported “not at all” for feeling down, depressed, or hopeless. For poor appetite or overeating, most respondents, 154 (46.2%), experienced this on “several days.” Trouble sleeping or sleeping too much was reported on “several days” by 171 (51.4%). The majority, 189 (56.8%), reported “not at all” for feeling bad about themselves or feeling like a failure. Similarly, 232 (69.7%) reported “not at all” for moving or speaking so slowly that others could have noticed or being restless. Regarding trouble concentrating, 154 (46.2%) reported experiencing it “not at all.” For thoughts of self-harm or being better off dead, the majority, 225 (67.6%), reported “not at all.”

**Table 3:** Assessment of depression among respondents.

| **Variables** | **Not at all**<br>**n(%)** | **Several days**<br>**n(%)** | **More than**<br>**half the**<br>**days**<br>**n(%)** | **Everyday**<br>**n(%)** |
| --- | --- | --- | --- | --- |
| Had little interest or pleasure in doing things | 168(50.5) | 131(39.3) | 28(8.4) | 6(1.8) |
| Had a poor appetite or been overeating | 141(42.3) | 154(46.2) | 34(10.2) | 4(1.2) |
| Feeling down, depressed, or hopeless | 168(50.5) | 119(35.7) | 37(11.1) | 9(2.7) |
| Had trouble falling or staying asleep, or been sleeping too much | 111(33.3) | 171(51.4) | 44(13.2) | 7(2.1) |
| Feeling tired or having little energy | 131(39.3) | 140(42.0) | 51(15.3) | 11(3.3) |
| Feeling bad about yourself - or that you are a failure or have let yourself or your family down | 189(56.8) | 119(35.7) | 21(6.3) | 4(1.2) |
| Moving or speaking so slowly that other people could have noticed or been restless | 232(69.7) | 73(21.9) | 18(5.4) | 10(3.0) |
| Had trouble concentrating on things, such as reading the newspaper or watching television | 154(46.2) | 140(42.0) | 28(8.4) | 11(3.3) |
| Had thoughts that you would be better off dead, or of hurting yourself in some way | 225(67.6) | 74(22.2) | 27(8.1) | 7(2.1) |

Figure 1 shows the different categories of depression among respondents. The majority (122, 65.5%) of respondents had mild depression, 46 (24.7%) had moderate depression while 18(9.7%) had moderately severe depression.

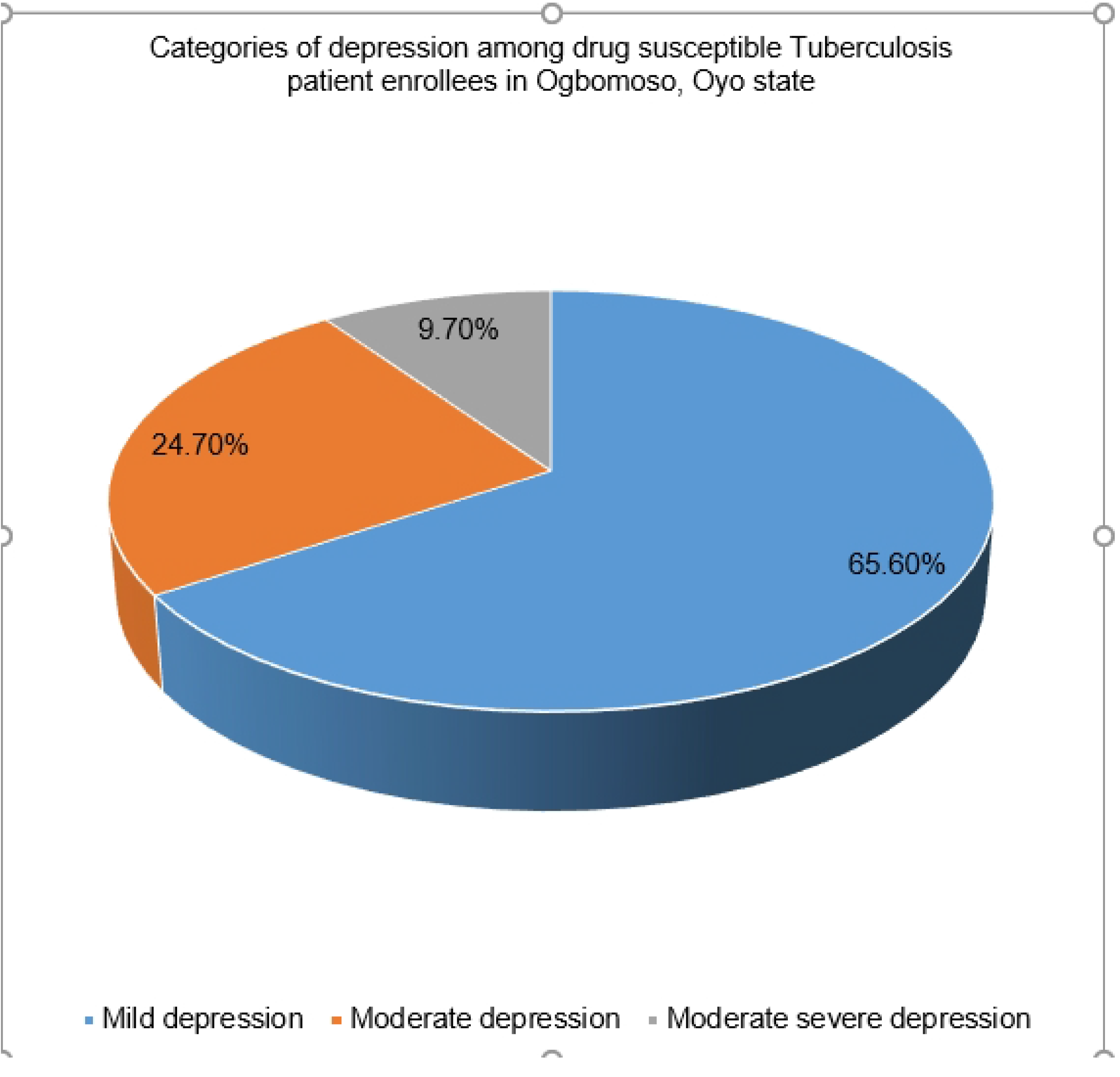

### Association between socio-demographic characteristics and depression status

Table 4 shows the relationship between socio-demographic characteristics and depression status revealed that sex (χ^2^ = 6.633, P = 0.010), marital status (χ^2^ = 23.948, P = <0.001), level of education (χ^2^ = 11.810, P = 0.008) and average monthly income (χ^2^ = 12.710, P = <0.001) were significantly associated with depression status among drug susceptible tuberculosis patient enrollees in Ogbomoso, Oyo state.

**TABLE 4:** ASSOCIATION BETWEEN SOCIO-DEMOGRAPHIC STATUS AND DEPRESSION STATUS.

| Variables | Categories | Depression | | $\chi^2$ | df | P |
| --- | --- | --- | --- | --- | --- | --- |
|  |  | Not depressed | Depressed |  |  |  |
| **Age Group** | <30 years | 14(10.3) | 14(9.6) | 1.230 | 4 | 0.873 |
|  | 30-40 years | 44(32.4) | 52(35.6) |  |  |  |
|  | 41-50 years | 38(27.9) | 33(22.6) |  |  |  |
|  | 51-60 years | 22(16.2) | 26(17.8) |  |  |  |
|  | >60 years | 18(13.2) | 21(14.4) |  |  |  |
| **Sex** | Male | 72(52.9) | 55(37.7) | 6.633 | 1 | **0.010\*** |
|  | Female | 64(47.1) | 91(62.3) |  |  |  |
| **Religion** | Christian | 71(52.2) | 82(56.2) | .581 | 2 | 0.748 |
|  | Muslim | 61(44.9) | 59(40.4) |  |  |  |
|  | Traditional | 4(2.9) | 5(3.4) |  |  |  |
| **Marital status** | Single | 17(12.5) | 43(29.5) | 23.948 | 3 | **<0.001\*** |
|  | Married | 108(79.4) | 78(53.4) |  |  |  |
|  | Widow | 53.7 () | 19(13.0) |  |  |  |
|  | Widower | 6(4.4) | 6(4.1) |  |  |  |
| **Ethnicity** | Yoruba | 111(81.6) | 115(78.8) | 6.904 | 3 | 0.075 |
|  | Igbo | 4(2.9) | 15(10.3) |  |  |  |
|  | Hausa | 16(11.8) | 13(8.9) |  |  |  |
|  | Others | 5(3.7) | 3(2.1) |  |  |  |
| **Occupational status** | Employed | 95(69.9) | 91(62.3) | 3.498 | 2 | 0.174 |
|  | Unemployed | 38(27.9) | 46(31.5) |  |  |  |
|  | Student | 3(2.2) | 9(6.2) |  |  |  |
| **Average income per month** | Less than ₦30 000 | 38(27.9) | 71(48.6) | 12.710 | 1 | **<0.001** |
|  | Above ₦30 000 | 98(72.1) | 75(51.4) |  |  |  |
| **Level of education** | No formal education | 28(20.6) | 23(15.8) | 11.810 | 3 | **0.008\*** |
|  | Primary | 45(33.1) | 32(21.9) |  |  |  |
|  | Secondary | 33(24.3) | 32(21.9) |  |  |  |
|  | Tertiary | 30(22.1) | 59(40.4) |  |  |  |

$\chi^2$ -Pearson chi square value, df-degree of freedom, P-Probability value, \*-significant at $P<.050$

### Association of HIV Status and Self-Reported Hypertension and Diabetes Status with Depression

Table 5 shows the relationship between the presence of comorbidities and the depression status of the respondents revealed that HIV status (χ^2^ = 61.381, P = <0.001) and Diabetes (χ^2^ = 7.295, P = 0.026) were significantly associated with depression status among drug susceptible tuberculosis patient enrollees in Ogbomoso, Oyo state.

**TABLE 5:** ASSOCIATION OF HIV STATUS AND SELF-REPORTED HYPERTENSION AND DIABETES STATUS WITH DEPRESSION.

| Variables | Categories | Depression | | Total | $\chi^2$ | df | P-value |
| --- | --- | --- | --- | --- | --- | --- | --- |
|  |  | Not depressed | Depressed |  |  |  |  |
| **HIV** | HIV positive | 3(2.2) | 60(41.1) | 63(22.3) | 61.381 | 1 | <0.001* |
|  | HIV negative | 133(97.8) | 86(58.9) | 219(77.7) |  |  |  |
| **Diabetes** | Yes | 19(14.0) | 24(16.4) | 43(15.2) | 7.295 | 2 | 0.026* |
| **Hypertension** | Yes | 29(21.3) | 36(24.7) | 65(23.0) | 4.446 | 2 | 0.108 |

$\chi^2$ -Pearson chi square value, df-degree of freedom, P -Probability value, \*-significant at $P < .050$

Table 6 above showing the predictors of depression status among the respondents revealed that respondents with no formal education were 6 times less likely to develop depression (AOR = 0.175, P = 0.001). Respondents with Primary level of education were 2 times less likely to develop depression (AOR = 0.427, P = 0.023). Respondents with Secondary level of education were 3 times less likely to develop depression compared to Tertiary level of education. Respondents living with HIV were 35 times more likely to develop depression (AOR = 35.303, P = 0.017) compared to those who were HIV negative.

**TABLE 6:**
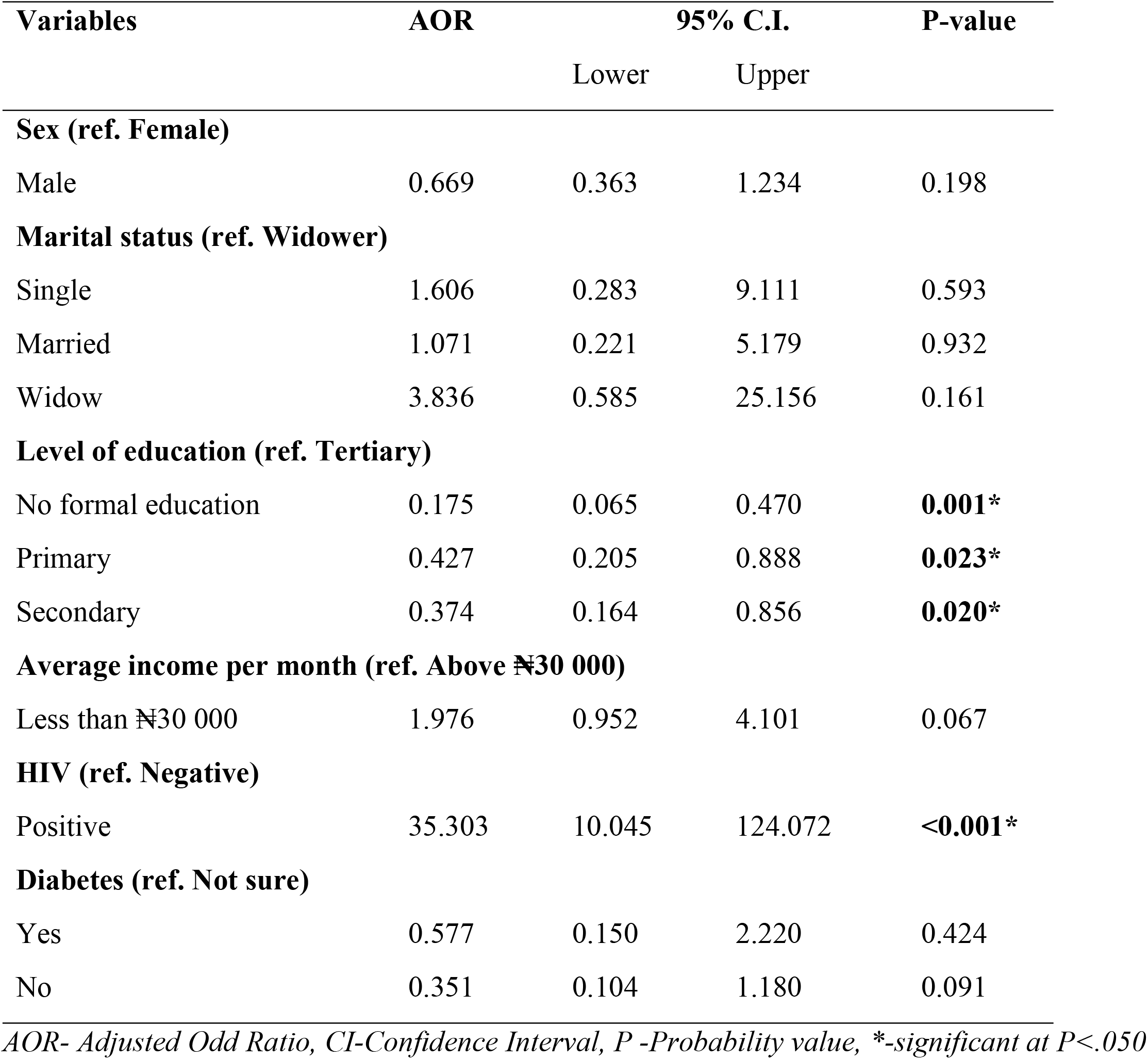
PREDICTORS OF DEPRESSION STATUS AMONG DRUG SUSCEPTIBLE TUBERCULOSIS PATIENT ENROLLEES IN OGBOMOSO, OYO STATE.

## DISCUSSION

The prevalence of depression among TB patients in this study was 51.8% which is found to be similar to the prevalence of 45.5%,^14^ 51.9%,^15^ 52.1%,^16^55.9%,^1^ and 54.0%,^17^ respectively from studies in Ethiopia, 45.5% in Southwest Nigeria,^13^ and considerably lower to studies in South Africa (64.3%),^18^ and Cameroon (61.1%).^2^ The observed differences could be due to the difference in population characteristics, prevalence among MDR-TB, time of assessment or phase of TB treatment.

This study observed that having no formal education, sex and marital status are statistically associated with depression, however, in contrast to many studies who have identified no formal education as a major predictor of depression,^3,5^ patients with no formal education in this study were observed to be six times less likely to develop depression. This could be due to lack of access to DOTS or poor health seeking behaviour resulting in a reduced population among TB patients receiving treatment. Being a male which was statistically associated with depression is however, not identified as a predictor in the multivariate analysis, similar to other studies who have identified the female gender as a significant predictor. This could be as a result of the global prevalence of depression among women,^16^ and other factors, including hormonal factors, household responsibilities and the social roles of women in the society have been mentioned.^2,16^ There was no association between age, religion, ethnicity and depression among the respondents in this study, however, some studies report old age as a significant predictor of depression among TB patients.^9,16^ Patients in the older age are believed to be prone to depression due to low financial status and susceptibility to TB stigma and the side effects of anti-TB drugs.^13^

Low socio-economic status among drug-susceptible TB patients was also found to be statistically associated with depression as seen in some other studies conducted among TB patients.^3,13,15^

Patients who received an average income less than the minimum wage were observed to be statistically associated with depression, however it was not identified as a predictor in the multivariate analysis.

The presence of TB/HIV comorbidity and diabetes mellitus is found to be statistically associated with depression in this study, which is consistent with other studies,^5,16^ and the TB-HIV coinfection may as a result of the stigma associated with HIV-positive patients, the depression associated with HIV infection.^16^ Patients living with TB/HIV comorbidity were observed to be 35 times likely to develop depression. This is considered significantly higher than other studies that observed the prevalence of depression in TB/HIV coinfection.^14^

## Data Availability

The data for this study is available on reasonable request from the corresponding author.

## ACKNOWLEDGEMENTS

The authors acknowledge the support of Professor Parakoyi, the laboratory scientists, research assistants and others who contributed to the success of the project.

## ETHICAL CONSIDERATION

Ethical approval for the study was obtained from the LAUTECH Teaching Hospital Ethical Review Committee and permission to carry out the study was obtained from the State Coordinator, Tuberculosis and Leprosy Control, Oyo State and from the Tuberculosis and Leprosy Control Supervisors in the chosen Local Government Areas.

Right of decline/withdraw from study: Respondents were told that participation is voluntary and they will not suffer any consequences if they chose not to participate.

Confidentiality of data: All information gathered were kept confidential and participants were identified using serial numbers.

Consent form: A written consent form signed by each respondent was included in the questionnaires.

Non-maleficence: No harm is intended nor befell any respondent in the course of the research study. Respondents were reassured of this.

Categories of depression among drug susceptible Tuberculosis patient enrollees in Ogbomoso, Oyo state

